# Confirmation of an Inverse Relationship between Bioaerosol Count and Influenza-like Illnesses, Including COVID-19. On the Contribution of Mold Spores

**DOI:** 10.1101/2021.02.07.21251322

**Authors:** Richa B. Shah, Rachna D. Shah, Damien G. Retzinger, Andrew C. Retzinger, Deborah A. Retzinger, Gregory S. Retzinger

## Abstract

Data from Chicago confirm the end of flu season coincides with the beginning of pollen season. The end of flu season also coincides with onset of seasonal aerosolization of mold spores. Overall, the data suggest bioaerosols, especially mold spores, compete with viruses for a shared receptor, with the periodicity of influenza-like illnesses, including COVID-19, a consequence of seasonal factors that influence aerosolization of competing species.

## Introduction

Influenza-like illnesses (ILIs) attributable to influenza viruses and to coronaviruses are sharply seasonal.^1,2^ Importantly, recent data from The Netherlands indicate there exists an inverse relationship between the seasonal incidence of influenza-like illnesses, including COVID-19, and pollen count.^3,4^ To discern whether such a relationship might be the case generally, pollen count in Chicago was related to ILIs reported by local emergency departments. In Chicago, as in The Netherlands, ILIs fall as total pollen count rises.

Because bioaerosols measured in Chicago include not only pollens but also mold spores, ILIs were related to counts of both. Just as they do for pollens, ILIs fall as mold spore count rises. *In contradistinction to their temporal relationship with pollens, however, ILIs remain low when mold spore count is high, rising again when mold spore count falls*.

Perusal of the various measured pollens and mold spores reveals many are echinulated, having protuberances reminiscent of those of influenza and SARS-CoV-2 viruses.^5–11^ Such protuberances seem ideally suited to interacting with Toll-like receptor 4 (TLR4) in a fashion akin to ‘hook-and-loop’ adhesives. It is proposed that the various pollens and mold spores compete with relevant respiratory viruses for TLR4, limiting viral engagement and, consequently, ILIs.

## Results

### Kinetics of Presentations to Emergency Departments

As shown in **Fig. 1A**, ILI presentations to emergency departments in Chicago are cyclical in nature, with a periodicity of ~ 1 year. The peak incidence occurs during ~ February, with the annual nadir occurring during ~ August. Over the entirety of a seasonal cycle, ILI presentations never reach zero. Although there are subtleties associated with the kinetics of ILIs for each of the individual years 2015 through 2020, the annual increase in ILI presentations for each season is characterized by a leading ‘bump,’ followed thereafter by a major rise. The relevance of the bump is addressed below, under both **Results** and **Discussion**. As for the major rise, it is approximated empirically by a first-order process, ILI_obs_ = ILI_0_e^kt^ + C, where ILI_obs_ is the number of observed ILIs during the growth phase of a cycle; ILI_0_ is the number of ILI presentations at the start of a cycle; t is time, in *d*; k is a first-order rate constant, in *d*^-1^; and C is the number of background presentations ‘masquerading’ as ILIs, **Table 1** and **Fig. 2A**. The rate of decrease from any yearly maximum also fits reasonably well a first-order process, **Table 1** and **Fig. 2B**. Taken at face value, the data suggest: 1) a not insignificant number of ILI presentations are not flu, and 2) the annual rate of change in flu cases is due to change in ambient concentration of influenza virus.

**Table 1.** Kinetics of ILI presentations to emergency departments in Chicago. Both the rate of appearance and the rate of disappearance of ILI presentations to Chicago emergency departments are approximately first-order. Using the boundaries indicated, data were fit to a first-order rate equation, which, for each year, was then solved for the parameters given in the table. See text for additional details.

Appearance
| Year | Date of Assigned Min. | Date of Assigned Max. | A<sub>0</sub>, Presentations (95%) | k, d<sup>-1</sup> (95%) | Half-life, d | C, Presentations (95%) | R<sup>2</sup> |
| --- | --- | --- | --- | --- | --- | --- | --- |
| 2014 | 1/9/2015 | 3/30/2015 | 5 (3 - 7) | 0.033 (0.028 - 0.042) | 21 | 36 (33 - 39) | 0.761 |
| 2015 | 10/27/2015 | 2/22/2016 | 1 (1 - 2) | 0.035 (0.029 - 0.043) | 20 | 40 (39 - 42) | 0.699 |
| 2016 | 10/22/2016 | 2/19/2017 | 2 (1 - 6) | 0.029 (0.022 - 0.038) | 24 | 35 (28 - 40) | 0.747 |
| 2017 | 10/9/2017 | 1/28/2018 | 19 (12 - 22) | 0.021 (0.019 - 0.025) | 34 | 0 (0 - 11) | 0.882 |
| 2018<sup>2</sup> | 10/28/2018 | 3/20/2019 | 39 (36 - 42) | 0.004 (0.003 - 0.005) | 185 | 0 (0 - 0) | 0.346 |
| 2019 | 11/11/2019 | 1/27/2020 | 49 (41 - 56) | 0.016 (0.014 - 0.019) | 43 | 0 (0 - 0) | 0.673 |
| 2020 | - | - | - | - | - | - | - |
| **Median** |  |  | **12** | **0.025** | **29** | **18** |  |

Disappearance
| Year | Date of Assigned Max. | Date Of Assigned Min. | A<sub>0</sub>, Presentations (95%) | k, d<sup>-1</sup> (95%) | Half-life, d | C, Presentations (95%) | R<sup>2</sup> |
| --- | --- | --- | --- | --- | --- | --- | --- |
| 2014 | - | - | - | - | - | - | - |
| 2015 | 3/30/2015 | 8/4/2015 | 66 (61 - 72) | 0.041 (0.034 - 0.049) | 17 | 29 (26 - 31) | 0.820 |
| 2016 | 2/22/2016 | 8/4/2016 | 113 (107 - 118) | 0.031 (0.027 - 0.035) | 22 | 18 (16 - 21) | 0.899 |
| 2017 | 2/19/2017 | 9/3/2017 | 105 (99 - 110) | 0.019 (0.017 - 0.022) | 36 | 13 (9 - 16) | 0.881 |
| 2018 | 1/28/2018 | 8/3/2018 | 155 (148 - 161) | 0.028 (0.026 - 0.031) | 24 | 20 (18 - 23) | 0.935 |
| 2019<sup>2</sup> | 3/20/2019 | 8/20/2019 | 63 (59 - 68) | 0.024 (0.020 - 0.029) | 29 | 16 (13 - 19) | 0.839 |
| 2020 | 3/18/2020 | 7/18/2020 | 145 (136 - 154) | 0.026 (0.021 - 0.030) | 27 | 10 (1 - 7) | 0.897 |
| **Median** |  |  | **109** | **0.027** | **26** | **17** |  |

**Figure 1.**
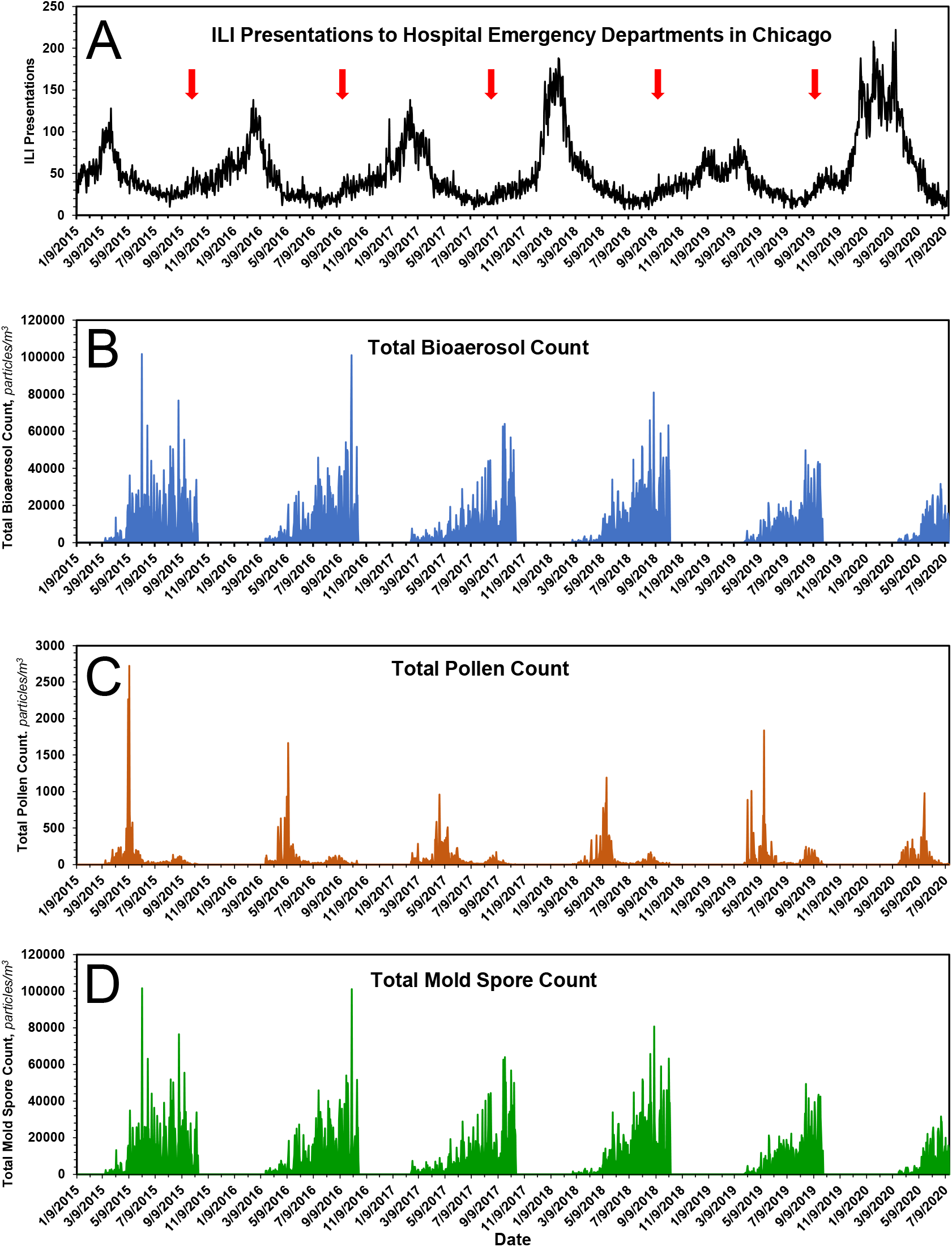
Seasonal time courses, 2015 – 2020, of measured variables of this study. *A*, ILI presentations to emergency departments of hospitals in Chicago. Red arrow indicates the leading seasonal ‘bump’ in ILI presentations. *B*, Total bioaerosol, *i*.*e*., pollens and mold spores, count as a function of time. *C*, Total pollen count as a function of time. *D*, Total mold spore count as a function of time. See text for additional details.

**Figure 2.**
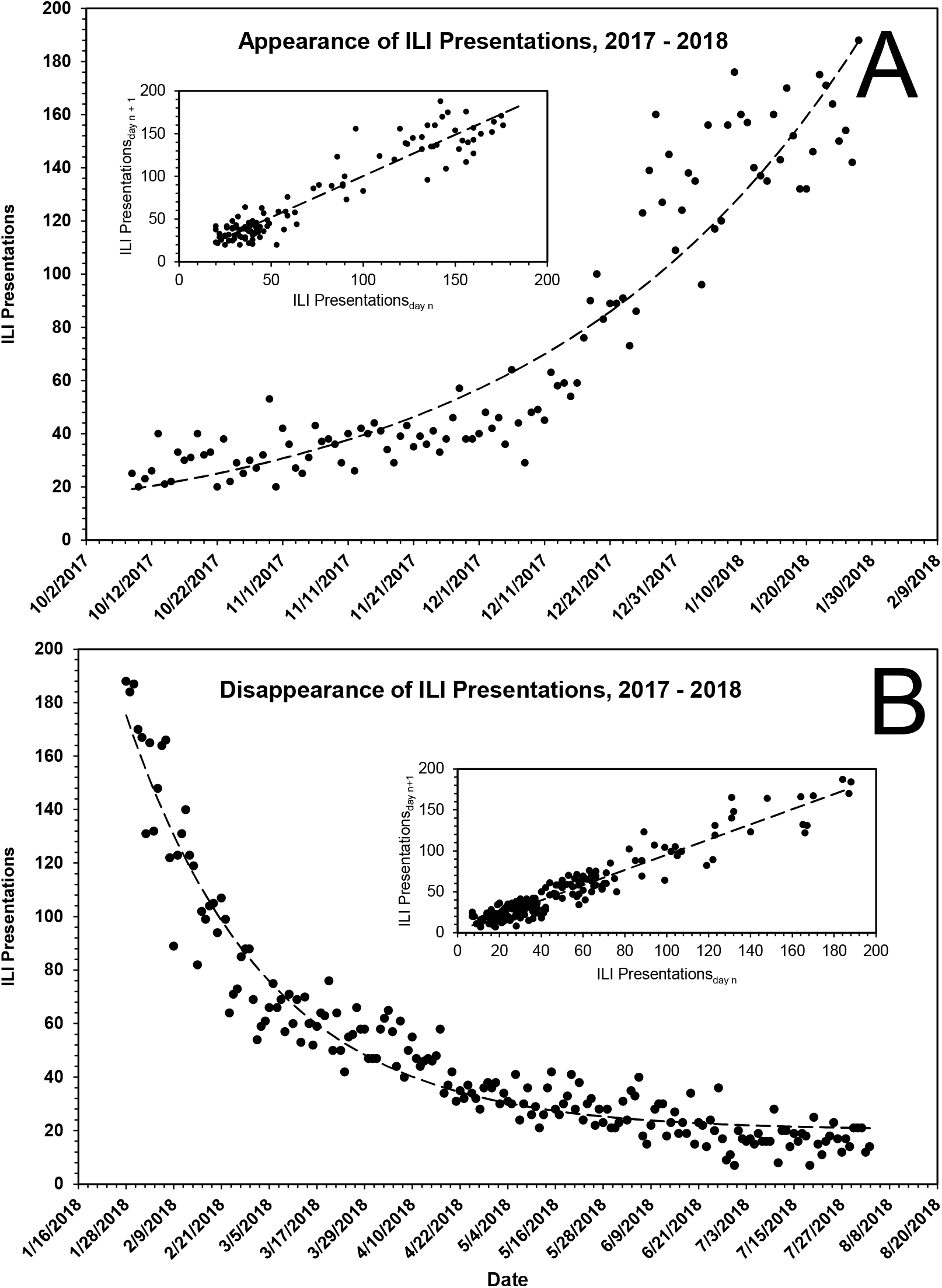
Kinetics of ILI presentations to hospitals in Chicago during a representative flu season, 2017 – 2018. Both the rate of appearance, *A*, and the rate of disappearance, *B*, of ILI presentations to emergency departments of Chicago hospitals are approximately first-order. The dashed lines displayed in the primary plots are the theoretical fits of the data (**Fig. 1A**) to a first-order rate equation, the parameters of which are given in **Table 2**. The inset shows the expected linearity of the same data when plotted according to the method of Kézdy,^12^ in this case *ILI presentations*_*day n*_ vs. *ILI presentations*_*day n+1*_.

**Fig. 3A** shows the time course of the 7-day moving average of COVID-19 presentations to emergency departments of all Chicago hospitals. Because reporting was not uniformly rigorous before May 1, 2020, COVID-19 presentations prior to that date have been excluded from analyses. As shown in **Fig. 3B**, the fall in presentations for the period May 5, 2020 through September 27, 2020 was roughly first-order, with parameters k = 0.065 (0.055 – 0.077) *d*^-1^ (t_1/2_ ~ 10.6 [9 – 13] *d*), COVID_0_ = 875 (799 – 954) *presentations* and C = 262 (241 – 281) *presentations*.

**Table 2.** Contribution of pollens and mold spores to bioaerosol burden in Chicago. The pollens and mold spores of this study contribute variously to bioaerosol burden in Chicago. Pollens are highlighted in yellow. *Particle Count* is the number of pollen or mold spore particles counted over the period of the study, ~ 6 years. *% Total* is the percentage of all the particles counted.

Bioaerosol Count, March 2015 – October 2020
| Bioaerosol | Particle Count* | % Total |
| --- | --- | --- |
| *Cladosporium* | 6064682 | 47.287 |
| Undifferentiated Ascospores | 3598475 | 28.058 |
| *Smuts / Myxomycetes* | 820505 | 6.398 |
| *Coprinus*-type | 600774 | 4.684 |
| *Ganoderma* | 368568 | 2.874 |
| *Alternaria* | 224356 | 1.749 |
| *Cercospora* | 208950 | 1.629 |
| *Diatrypaceae* | 116333 | 0.907 |
| *Penicillium / Aspergillus* | 109272 | 0.852 |
| *Epicoccum* | 97962 | 0.764 |
| Undifferentiated Basidiospores | 93886 | 0.732 |
| Rusts | 93107 | 0.726 |
| *Drehslera / Helminthosporium* | 79746 | 0.622 |
| *Pithomyces* | 45102 | 0.352 |
| *Leptosphaeria*-type | 40322 | 0.314 |
| *Nigrospora* | 36701 | 0.286 |
| *Torula* | 32652 | 0.255 |
| ***Morus*** | **28019** | **0.218** |
| *Periconia* | 26389 | 0.206 |
| *Chaetomium* | 14408 | 0.112 |
| *Curvularia* | 14166 | 0.110 |
| *Oidium / Erysiphe* | 12012 | 0.094 |
| *Stemphylium* | 11304 | 0.088 |
| *Fusarium* | 10277 | 0.080 |
| *Polythrincium* | 9714 | 0.076 |
| ***Populus*** | **8034** | **0.063** |
| ***Quercus*** | **6792** | **0.053** |
| ***Acer*** | **6576** | **0.051** |
| ***Ambrosia*** | **5896** | **0.046** |
| ***Gramineae / Poaceae*** | **5399** | **0.042** |
| *Peronospora* | 4676 | 0.036 |
| ***Chenopodiaceae/Amaranthaceae*** | **4637** | **0.036** |
| ***Cupressaceae*** | **4395** | **0.034** |
| ***Betula*** | **4132** | **0.032** |
| ***Pinaceae*** | **2607** | **0.020** |
| Unidentified Fungi | 2366 | 0.018 |
| ***Urticaceae*** | **2337** | **0.018** |
| ***Plantago*** | **1584** | **0.012** |
| *Pleospora* | 1473 | 0.011 |
| ***Salix*** | **1284** | **0.010** |
| ***Artemisia*** | **1019** | **0.008** |
| ***Fraxinus*** | **823** | **0.006** |
| ***Ulmus*** | **822** | **0.006** |
| Unidentified Pollen | 699 | 0.005 |
| ***Rumex*** | **483** | **0.004** |
| ***Juglans*** | **386** | **0.003** |
| ***Alnus*** | **281** | **0.002** |
| ***Platanus*** | **231** | **0.002** |
| ***Tilia*** | **170** | **0.001** |
| ***Carya*** | **147** | **0.001** |
| ***Cyperaceae*** | **128** | **0.001** |
| ***Fagus*** | **120** | **0.001** |
| ***Liquidambar*** | **85** | **0.001** |
| ***Typha*** | **38** | **0.000** |
| ***Asteraceae (excl. Ambrosia and Artemisia)*** | **38** | **0.000** |
| ***Olea*** | **18** | **0.000** |
| *Botrytis* | 0 | 0.000 |

\*Total across all seasons

**Figure 3.**
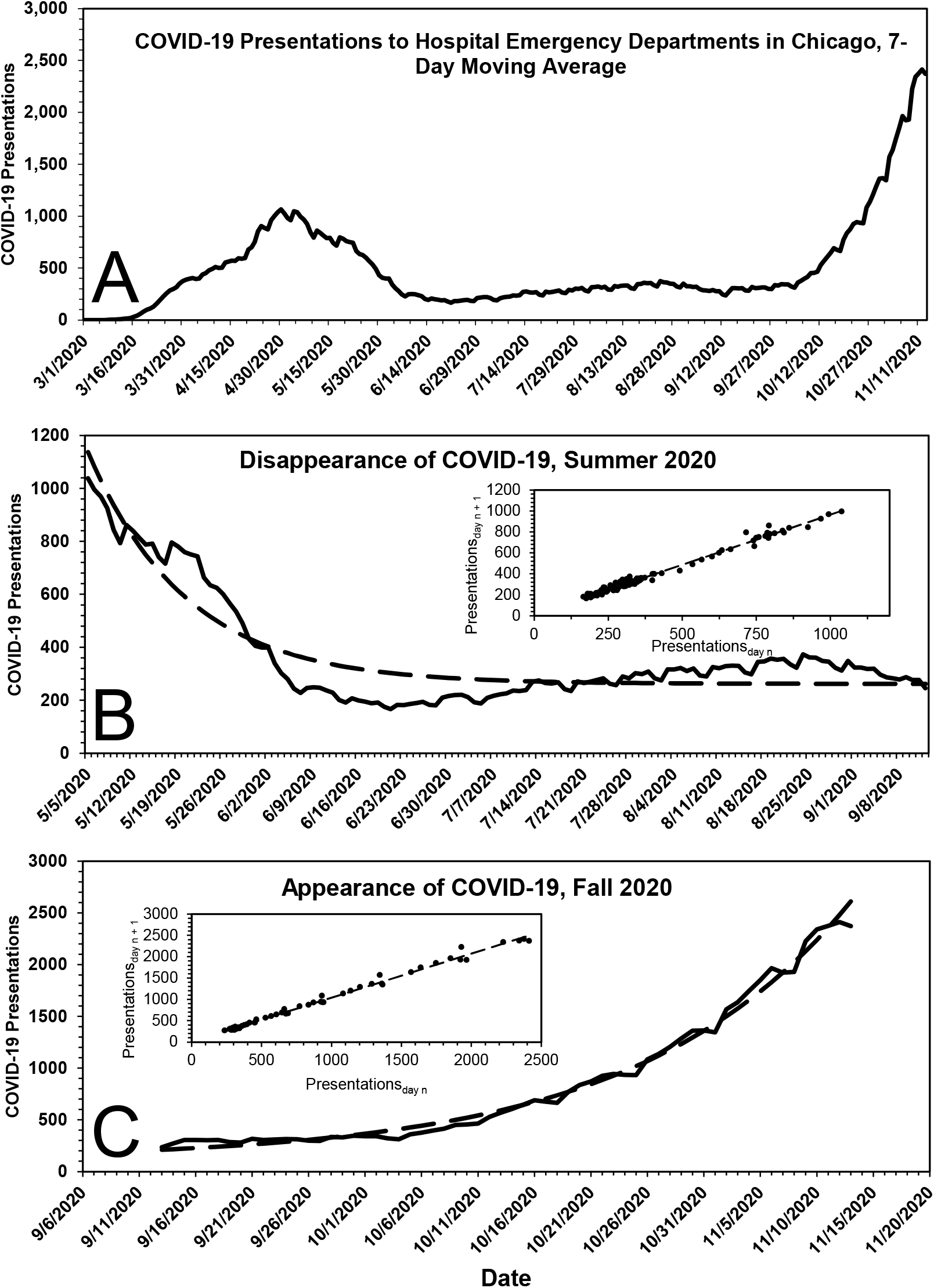
Time course and kinetics of COVID-19 presentations to all Chicago hospitals, March 2020 – November 2020. *A*, COVID-19 presentations to all hospitals in Chicago, 7-day moving average, March 1, 2020 to November 14, 2020. *B*, Disappearance of COVID-19 presentations to all hospitals in Chicago, May 5, 2020 to September 13, 2020. The dashed line displayed in the primary plot is the theoretical fit of the data to a first-order rate equation, the parameters of which are given in the text. The inset shows the expected linearity of the same data when plotted according to the method of Kézdy,^12^ in this case *COVID-19 presentations*_*day n*_ vs *COVID-19 presentations*_*day n+1*_. *C*, Appearance of COVID-19 presentations to all hospitals in Chicago, September 14, 2020 to November 14, 2020. The dashed line displayed in the primary plot is the theoretical fit of the data to a first-order rate equation, the parameters of which are given in the text. The inset shows the expected linearity of the same data when plotted according to the method of Kézdy,^12^ in this case *COVID-19 presentations*_*day n*_ vs. *COVID-19 presentations*_*day n+1*_.

Starting in late September 2020, COVID-19 cases in Chicago surged in first-order fashion, **Fig. 3C**, the parameters of which are k = 0.053 (0.050 – 0.057) *d*^-1^ (t_1/2_ ~ 13 [12– 14] *d*), COVID-19_0_ = 96 (76 – 119) *presentations* and C = 117 (63 – 165) *presentations*. Taken at face value, the data suggest: 1) some of the individuals for whom a diagnosis of COVID-19 was made did not have COVID-19, and 2) changes in the number of COVID-19 presentations in Chicago were due to changes in the ambient concentration of SARS-CoV-2.

### Kinetics of Pollen and Mold Spore Counts

Pollens are fertilizing elements of flowering plants whilst mold spores are reproductive elements of fungi. In published studies,^3,4^ pollens alone were counted and related to ILIs. Left uncounted were mold spores, important seasonal contributors to the total bioaerosol burden. For the studies reported herein, both pollens and mold spores were counted. Those counts were then analyzed, in aggregate and individually.

Usually, pollens and mold spores in Chicago are monitored from ~ mid-March to ~ mid-October, the period most problematic for persons suffering from seasonal allergies. As expected, the data indicate bioaerosol expression is cyclical with a periodicity of ~ 1 year, **Fig. 1B**. The total bioaerosol count peaks during ~ mid-September and falls sharply thereafter. Unfortunately, data following the peaks are somewhat limited, their collection being truncated on an arbitrary end date, *i*.*e*., ~ mid-October.

In the case of pollens, the seasonal distribution is bimodal, with a dominant first mode that peaks in ~ mid-May and a smaller second mode that peaks in ~ late August, **Fig. 1C**. The pollens that constitute the second mode, here termed ‘late pollens’, are predominantly *Ambrosia* and the other *Asteraceae*. Importantly, the peak of the second mode always coincides with the leading bump in ILI presentations, **Figs. 1A** and **4**. The potential relevance of this is addressed under **Discussion**. In the case of mold spores, which constitute the bulk of the measured bioaerosols, **Figs. 1B** and **1D**, and **Table 2**, the peak count, which occurs during ~ late September, falls precipitously by ~ mid-October, with an empiric half-life of ~ 10 *d*, **Table 3**.

**Table 3.** Kinetics of mold spore expression in Chicago. The rate of appearance of mold spores in Chicago is approximately first-order. Using the boundaries indicated, data were fit to a first-order rate equation, which, for each year, was then solved for the parameters given in the table. See text for additional details. Because the end date for counting pollens and mold spores in Chicago is fixed at ~ mid-October, the rate of disappearance of mold spores is approximated by the change in mold spore count over the interval of time indicated in the table.

Disappearance
| Year | Date of Assigned Max., A | Mold Spore Count on A,<br>*particles/m<sup>3</sup>* | Date of Last<br>Measurement, B | Mold Spore Count on B, *particles/m<sup>3</sup>* | Interval (A to B), *d* |
| --- | --- | --- | --- | --- | --- |
| 2015 | 9/2/2015 | 76403 | 10/16/2015 | 10344 | 44 |
| 2016 | 10/6/2016 | 101028 | 10/21/2016 | 7203 | 15 |
| 2017 | 9/25/2017 | 63929 | 10/20/2017 | 10519 | 25 |
| 2018 | 9/5/2018 | 80759 | 10/12/2018 | 22466 | 37 |
| 2019 | 9/20/2019 | 43414 | 9/30/2019 | 12920 | 10 |
| 2020 | 9/4/2020 | 57100 | 10/13/2020 | 5200 | 39 |
| **Median** |  | **70166** |  | **10432** | **31** |

Appearance
| Year | Date of First Observed<br>Appearance | Date of Assigned Max. | Interval, *d* | *M*<sub>0</sub>, *particles/m<sup>3</sup>*<br>(95%) | *k*, *d*<sup>-1</sup> (95%) | Half-life, *d* | *R*<sup>2</sup> |
| --- | --- | --- | --- | --- | --- | --- | --- |
| 2015 | 3/16/2015 | 9/2/2015 | 170 | 7350 (5427 - 9923) | 0.008 (0.006 - 0.011) | 84 | 0.211 |
| 2016 | 3/21/2016 | 10/6/2016 | 199 | 3759 (2788 - 5081) | 0.012 (0.010 - 0.014) | 59 | 0.478 |
| 2017 | 2/20/2017 | 9/25/2017 | 217 | 2682 (4418 - 7437) | 0.011 (0.006 - 0.009) | 65 | 0.395 |
| 2018 | 3/1/2018 | 9/5/2018 | 188 | 2256 (1489 - 3142) | 0.015 (0.013 - 0.018) | 45 | 0.572 |
| 2019 | 4/1/2019 | 9/24/2019 | 176 | 3137 (1399 - 3632) | 0.013 (0.012 - 0.019) | 52 | 0.634 |
| 2020 | 3/23/2020 | 9/4/2020 | 165 | 4044 (2820 - 5624) | 0.012 (0.010 - 0.015) | 56 | 0.479 |
| **Median** |  |  | **188** | **3137** | **0.012** | **59** |  |

Although these data substantiate the claim of an inverse relationship between the onset of pollen season and the end of flu season, pollen count declines rapidly and is not elevated when ILI presentations, **Fig. 5A**, and COVID-19 presentations, **Fig. 6A**, are low. Mold spores, on the other hand, increase continuously in first-order fashion, **Table 3** and **Fig. 7**, beginning just prior to or coincident with the fall in ILI, **Fig. 5B**, and COVID-19 presentations, **Fig. 6B**, and across the entirety of the summer months, when influenza and COVID-19 cases are low.

**Figure 5.**
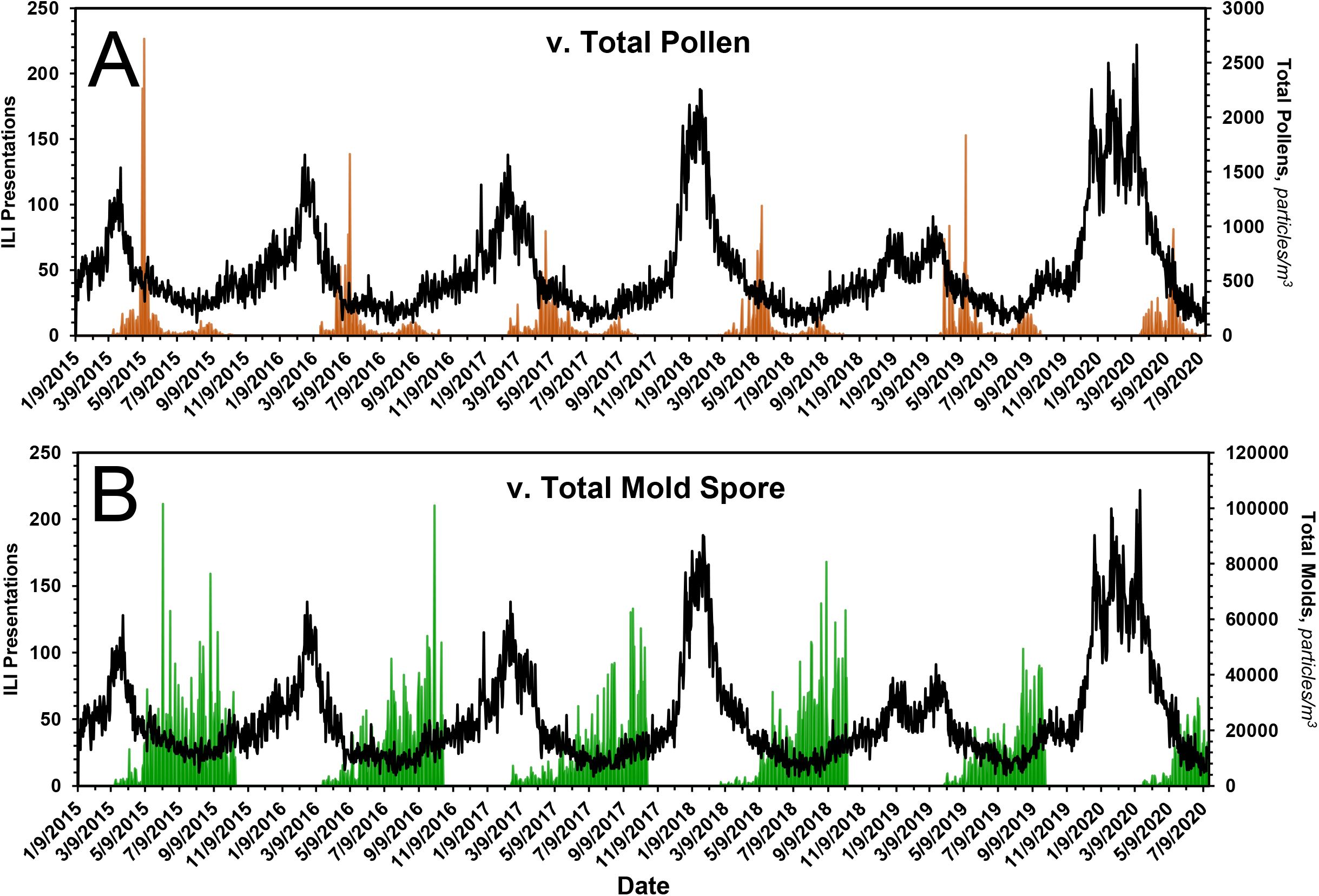
Bioaerosol expression and ILI presentations in Chicago, 2015 – 2020. In *A*, the expression of pollens, in brown, is superimposed on the time course of ILI presentations. In *B*, the expression of mold spores, in green, is superimposed on the time course of ILI presentations. The expression of pollens is bimodal, with the onset of the first mode coinciding with the drop in seasonal ILI presentations. The peak of the second mode coincides with the onset of the leading ‘bump’ in ILI presentations, the start of flu season. See text and **Fig. 4** for additional details. The onset of aerosolization of mold spores also coincides with the drop in seasonal ILI presentations. Thereafter, mold spore count increases across the entirety of the summer months --during which time ILIs remain low -- and falls precipitously from mid-September to mid-October, at which time ILI cases begin to rise. See text for additional details.

**Figure 6.**
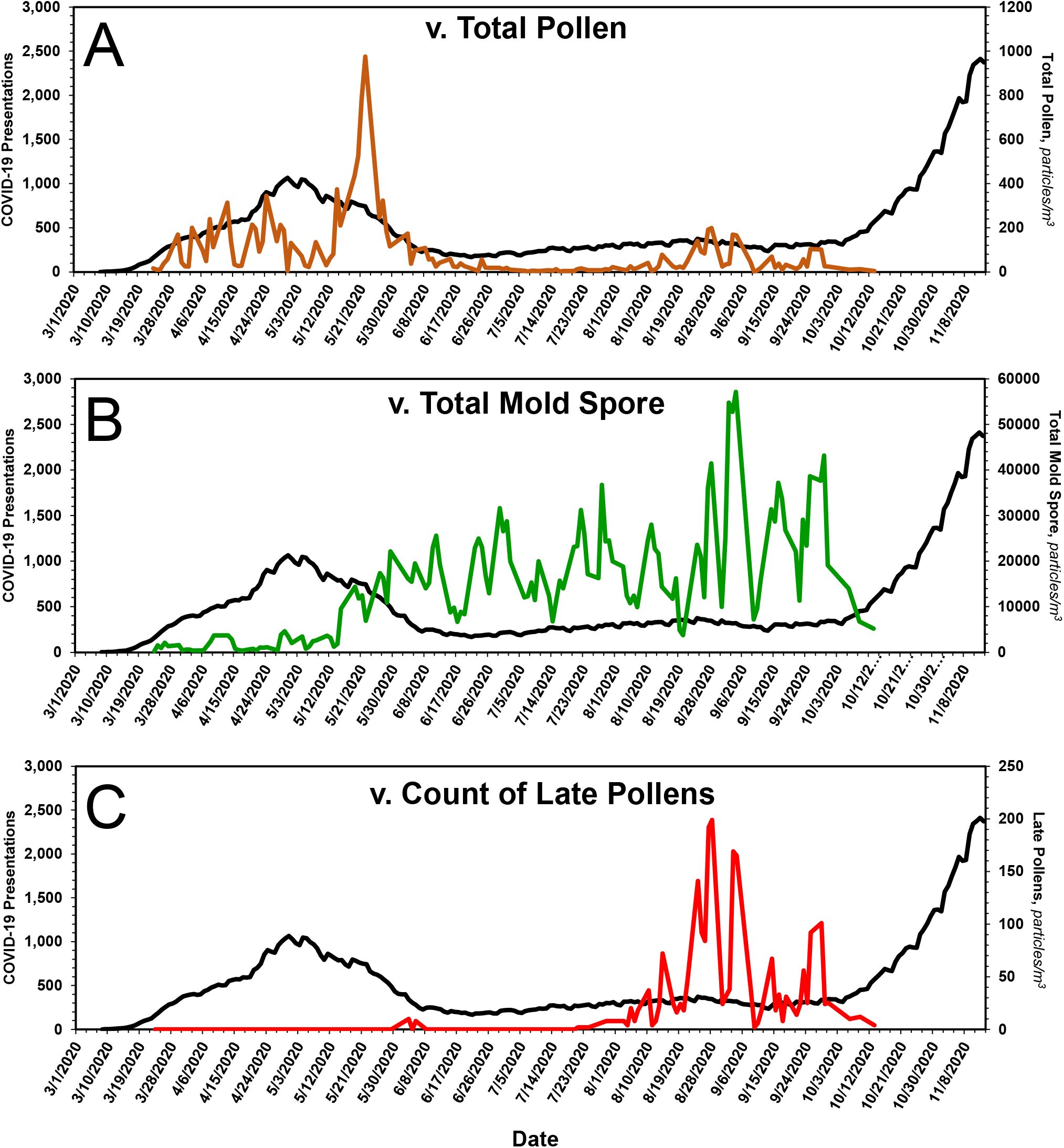
Bioaerosol expression and COVID-19 presentations in Chicago, March 2020 – November 2020. In *A*, the expression of pollens, in brown, is superimposed on the time course of COVID-19 presentations. In *B*, the expression of mold spores, in green, is superimposed on the time course of COVID-19 presentations. In C, the expression of late pollens alone, in red, is superimposed on the time course of COVID-19 presentations. As they do for ILI presentations, late pollens presage the onset of COVID-19 presentations in the Fall. The onset of aerosolization of mold spores coincides with the drop in COVID-19 presentations in the Spring. Thereafter, mold spore count increases across the entirety of the summer months --during which time COVID-19 presentations remain low -- and fall precipitously from mid-September to mid-October, at which time COVID-19 presentations begin to rise. See text for additional details.

**Figure 7.**
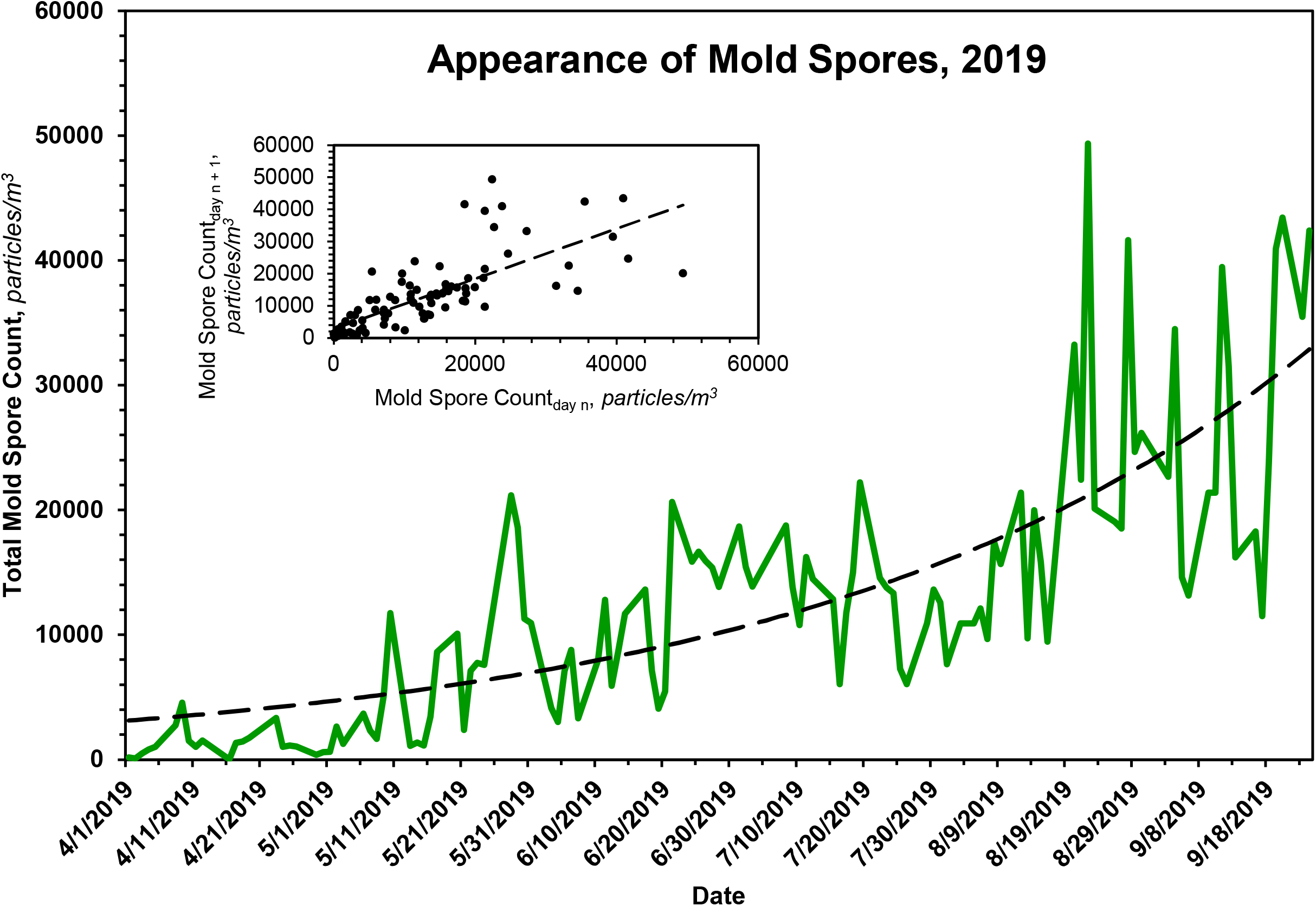
Kinetics of mold spore expression in Chicago during a representative season, 2019. The rate of appearance of mold spores in Chicago is approximately first-order. The dashed line displayed in the primary plot is the theoretical fit of the data to a first-order rate equation, the parameters of which are given in **Table 4**. The inset shows the expected linearity of the same data when plotted according to the method of Kézdy,^12^ in this case *mold spore count*_*day n*_ vs. *mold spore count*_*day n+1*_.

### Inhibition of ILI and COVID-19 Presentations by Mold Spores

If one assumes ILI and COVID-19 presentations are consequences of the binding of relevant viruses to specific receptors, then one can treat the presentations as proxies for those receptors, for which mold spores compete. Toward that end, ILI and COVID-19 presentations were plotted as functions of total mold spore count, **Fig. 8**. Because the curvatures of the plots suggest true equilibria, the data of each were fit to the equation P = P_o_/(1+C/K_d_) + B, where P is the observed number of presentations to emergency departments, P_o_ is the maximum number of such presentations, C, in *mold spores*/*m*^*3*^, is the measured mold spore count, K_d_, in *mold spores*/*m*^*3*^, is the apparent dissociation constant of the receptor – mold spore complex, and B is a constant representing presentations not influenced by mold spores. As shown in the figures, the data of each plot fit reasonably well the theoretical model. From the ILI data, one calculates P_o_ ~ 50 (41 – 60) *presentations*, K_d_ ~ 2128 (997 – 4242) *mold spores*/*m*^*3*^ and B ~ 16 (9 – 20) *presentations*; from the COVID-19 data, one calculates P_o_ ~ 1366 (994 – 2679) *presentations*, K_d_ ~ 1668 (487 – 4143) *mold spores*/*m*^*3*^.and B ~ 201 (103 – 257) *presentations*. The most parsimonious explanation for the near equivalence of the apparent dissociation constants is a shared receptor.

**Table 4.**
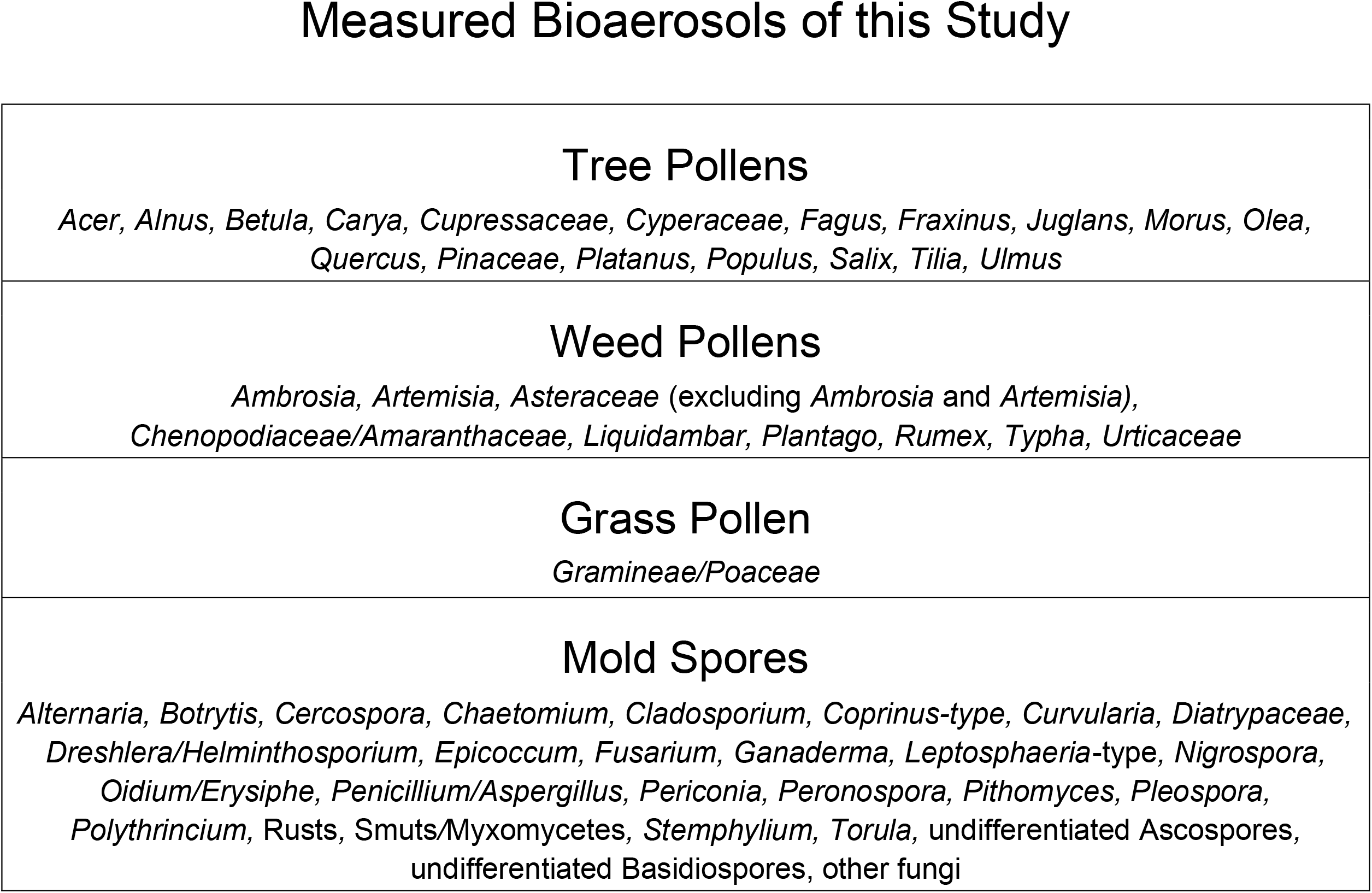
Bioaerosols of this study. The measured bioaerosols of this study, *i*.*e*., pollens and mold spores, are those listed here. They were collected and quantified as described in the text.

**Figure 8.**
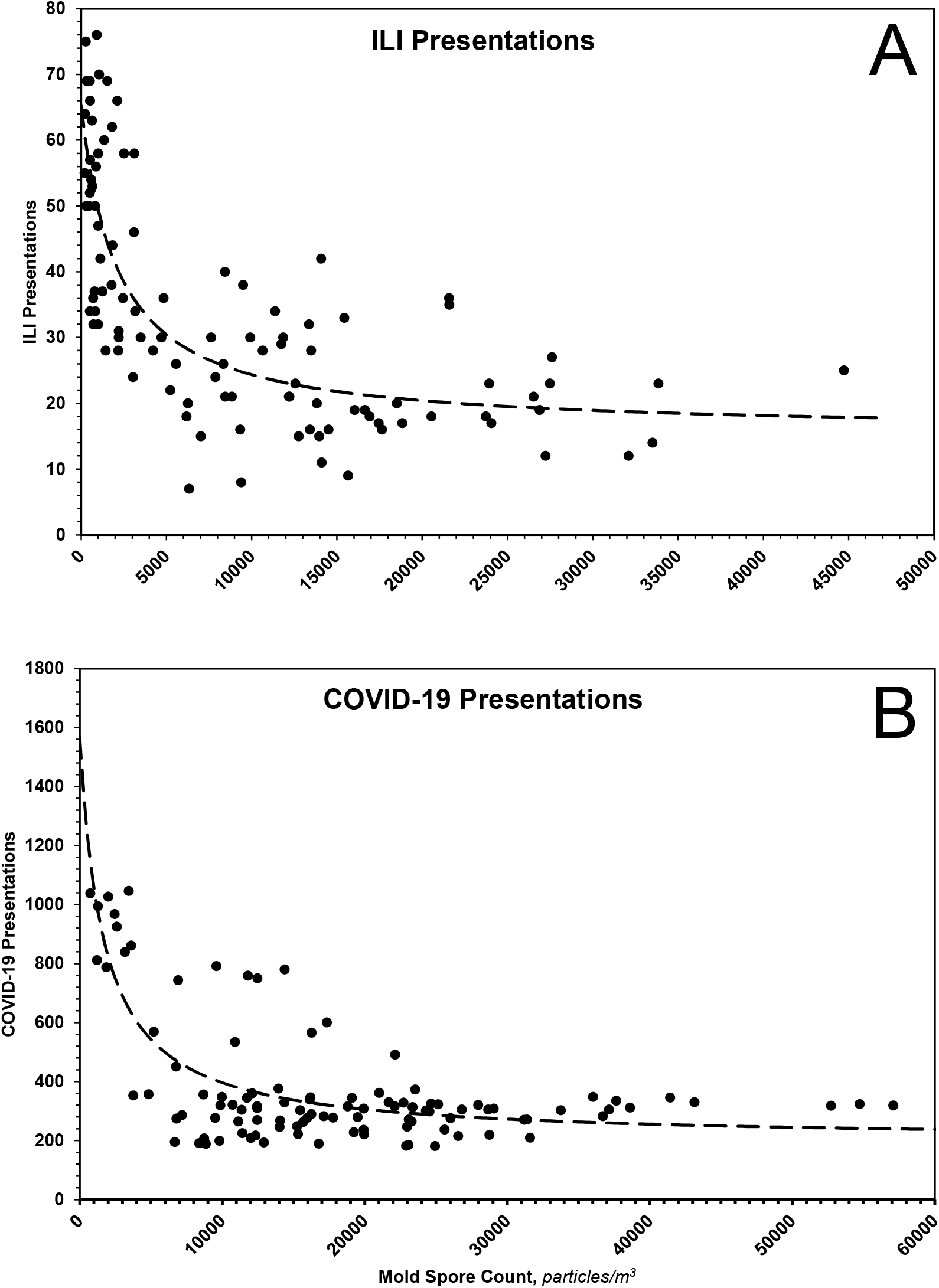
Inhibition of ILIs, A, and COVID-19, B, by mold spores. The dotted lines represent theoretical fits of data according to the equation and parameters given in the text.

## Discussion

The data presented herein are consistent with those presented earlier by others,^3,4^ namely, the incidence of ILIs falls as pollen count rises. Because the data of the present study derive from an urban area in North America (Chicago, IL USA: latitude 41.85003, longitude −87.65005) whilst those of the earlier study derive from North Central Europe (Helmond, The Netherlands: latitude 51.48167, longitude 5.66111), it appears the inverse relationship may be generally valid.

With special regard to the late pollens, changes in their atmospheric concentration invariably coincide with the annual leading bump in ILIs. Inasmuch as *Ambrosia*, the dominant species, is a major respiratory allergen, the leading bump may represent ragweed sensitivities manifesting as ILI. Regardless, the peak in late pollen count --as if a switch --presages the major upswing in ILI, **Fig. 4**, and COVID-19, **Fig. 6C**, presentations. Thus, aside from any contribution to mechanistic understanding it might provide, the peak in late pollens could be exploited when contemplating an upcoming ILI season.

**Figure 4.**
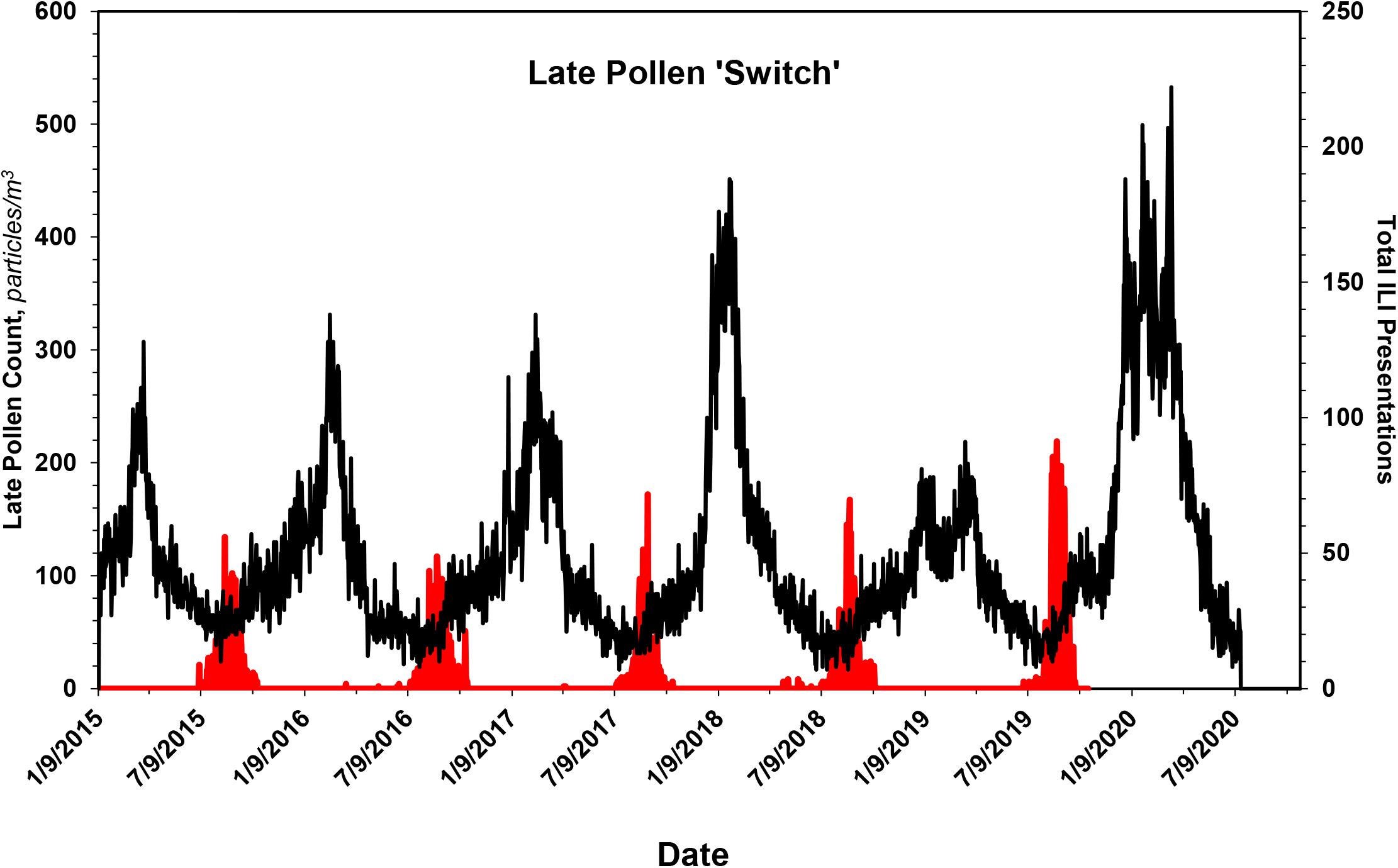
Late pollens signal the start of the flu season. Data shown in red are solely the counts of late pollens, most of which are *Ambrosia* and other *Asteraceae*. Superimposed on them are the ILI data of **Fig. 1A**. As if a switch, the peak in the count of late pollens always occurs coincident with onset of the bump in seasonal ILI presentations. See text for additional details.

Separate and distinct from pollen count, mold spore count in Chicago correlates inversely with ILIs. Indeed, given their higher atmospheric concentration as well as the duration of their seasonal expression, mold spores seem more likely than pollens to be principals in any abatement of ILIs, including COVID-19. Mold spores and pollens could abate viral activity by either direct or indirect means. By direct means, they might produce substances that limit viral propagation or they might complex with viruses, limiting viral infectivity.^13,14^ But if direct antiviral activity is an attribute of the bioaerosols themselves, then one would not expect, *a priori*, significant disparity between individual susceptibilities to severe flu or COVID-19.^15,16^ As indirect means, others have proposed pollens stimulate the human immune system in such a way as to either potentiate endogenous antiviral activity or elicit a protective allergic response.^3^ Against these proposals, asthma does not confer protection against either influenza or COVID-19.^17–19^

The similarity of the proposed mold spore dose dependencies for abatement of flu and COVID-19 suggests a shared receptor. Although much attention has been given to angiotensin-converting enzyme 2 (ACE-2) and its role in COVID-19,^20,21^ there are compelling reasons to believe TLR4, which binds the SARS-CoV-2 spike protein with greater affinity than does ACE-2,^22^ is also operative: 1) TLR4 is implicated in the inflammatory response triggered by sharply seasonal respiratory viruses,^23–25^ 2) TLR4 has a significant role in innate defense against multiple species of fungi,^26,27^ and polymorphisms in TLR4 are associated with invasive fungal disease,^28,29^ 3) COVID-19 prognosis correlates with radiographic involvement of alveolar spaces,^30,31^ the epithelial surfaces of which are poor in ACE-2^32,33^ but rich in TLR4,^34^ 4) inflammation of the sort associated with acute lung injury is mediated by TLR4,^35–44^ and 5) age-dependent hyper-responsiveness of TLR4^45^, especially in the context of interactions with TLR5,^46,47^ can account for the age-dependent severity of COVID-19. That TLR4 may be involved in the processing of bioaerosols is also expected on phylogenetic grounds: the receptor has been retained by some fish that breathe air, but lost by those that do not,^48^ and the eponymous Toll receptor controls the antifungal response of *Drosophila*.^49^

Given these, one can imagine the engagement of TLR4 by aerosols of all sorts, including, but not limited to, viruses, pollens and mold spores, in a fashion analogous to the engagement of hook-and-loop adhesives, *i*.*e*., Velcro^®^. Instead of loops, however, spinous processes of the various aerosols engage TLR4 ‘hooks,’ effecting an innate immune response, the nature of which depends on the arrangement and density of the engagement. And just as hook-and-loop adhesives can be rendered nonfunctional/dysfunctional by nonspecific adherence of extraneous materials, so, too, might TLR4 hooks become saturated with one ligand, *e*.*g*., mold spores and/or pollens, to the exclusion of another, *e*.*g*., a respiratory virus.

The data presented herein bring new appreciation and understanding to seasonality and suggest a remarkable interplay between bioaerosols that influence the health of man. Indeed, inasmuch as humans have co-existed with plants, fungi and viruses for some time, it stands to reason that, over the course of evolution, the respiratory system of the former would have developed means to cope with the significant recurring, *i*.*e*., annual, inhalational exposure to reproductive elements of the latter. As the environment-facing interface of the respiratory tree, epithelial cells and their entourage of innate immune effectors seem ideally positioned to provide that coping mechanism.

## Materials and Methods

### Collection and Counting of Pollens and Mold Spores

A volumetric spore trap (Burkard Manufacturing, Hertfordshire, England) equipped with a 24 *h* sampling head was used to collect pollens and mold spores. The trap was fixed ~ 70 feet above ground, on a roof in Melrose Park, Il, USA. A standard glass microscope slide coated with grease was placed in a carriage that moved at a rate of 2 *mm/h* past the trap orifice (14 *mm* x 2 *mm*). Air was drawn through the orifice at a rate of 10 *l/min*, thereby impacting airborne particles against the greased slide. Slides so exposed were stained with glycerin jelly supplemented with basic fuchsin. After applying a coverslip, a slide was evaluated microscopically for both pollens and mold spores, **Table 4**. A new slide was placed in the trap daily, and the carriage was re-oriented to its start position. Bioaerosol counts were made Monday through Friday, generally between mid-March and mid-October.

### ILI and COVID-19 Data

ILI data pooled from 23 large hospitals in Chicago over the period January 9, 2015 through July 18, 2020 were obtained from the Chicago Department of Public Health (CDPH). The 23 hospitals were chosen because they alone of Chicago-area hospitals consistently reported ILI presentations over the entirety of the study interval. The data are included in the *Supplement, Table S1*. COVID-19 data from all Chicago hospitals were obtained through portals of the CDPH, https://www.chicago.gov/city/en/sites/covid19/home/covid-dashboard.html and https://data.cityofchicago.org/browse?limitTo=datasets&sortBy=alpha&tags=covid-19. Those data are also included in the *Supplement, Table S2*.

## Data Availability

All data generated or analyzed during this study are included in this published article and its supplementary information files.

## Data Analysis

Time- or dose-dependent data were paired with the corresponding time or dose and fit to equations described in the text. The best values for the parameters of the equations, as well as their corresponding 95% confidence intervals, were then determined using the paired data and a nonlinear least squares regression method.^50^

## Acknowledgment

D.A.R. and G.S.R. thank their children, Andrew, Jonah, Ruth and Damien, for inspiration.

## Author Contributions

R.B.S. collated all the data, and contributed to preparing and writing the manuscript. R.D.S. collected the pollen and mold spore data, and contributed to writing the manuscript. D.G.R. processed and analyzed the data, and contributed to preparing and writing the manuscript. A.C.R. analyzed the data and contributed to preparing and writing the manuscript. D.A.R. conceived parts of the manuscript and contributed to its preparation and writing. G.S.R. conceived and designed the study, processed and analyzed the data, and contributed to preparing and writing the manuscript.

